# Sun exposure behavior and knowledge on photoprotection of children and adolescents living in tropical regions

**DOI:** 10.1101/2022.02.09.22270727

**Authors:** Marcelo de Paula Corrêa, Fabrina Bolzan Martins, Ana Letícia Campos Yamamoto, Natana Batista Paiva, Luiz Felipe Silva, Reynaldo José Sant’Anna Silva, Mayara Motta Melo, Clarissa Santos de Carvalho Ribeiro

## Abstract

**Background:** Excessive and continuous exposure to ultraviolet radiation (UVR) can cause skin cancer (SC), photoaging, and cataracts. Photoprotection (PhP) is the most important tool in preventing against these harmful effects. Elementary and high school students should be targeted in PhP campaigns because the solar radiation that youngers receive increases the risk of SC in adulthood. However, PhP policies are scarce in developing countries.

**Objectives:** To evaluate the use and knowledge of PhP with youngsters living in tropical regions.

**Methods:** 2,844 students aged from 4 to 20 years-old were interviewed about their knowledge on sun exposure and PhP habits. Age, sex, family income and skin phototype were statistically related to the proposed questions.

**Results:** Brazilian youngsters do not have the habit of wearing long-sleeved clothing (<15% in summer), nor sunglasses (0.0%), and hats (2.1%). More than 40% had no knowledge about UVI while another 30% misinterpret what it is. Less than half of all students wear sunscreen when out in the sun. Despite this low PhP use-rate, more than 90% know that UVR causes SC. Income was the factor that showed the greatest number of significant relationships respect to PhP.

**Conclusions:** Children and adolescents showed little knowledge about the sun exposure effects and lack PhP use. The lack of educational programs predisposes these youngsters to an increased chance of SC in the future. These results suggest that PhP campaigns should be part of school curricula from elementary school onwards.

## 1. Introduction

Skin cancer (SC) is a public health problem worldwide, especially in tropical and subtropical regions, due to the high incidence of ultraviolet solar radiation (UVR) in these areas [1,2]. The number of cases of SC has increased worldwide in recent years, especially in its less common yet more severe form, cutaneous melanoma (CM). According to the World Health Organization, more than 130,000 new cases of CM occur globally each year [3]. The most common SCs are non-melanoma skin cancers (NMSC), called basal cell carcinomas (BCC), and squamous cell carcinomas (SCC), constituting 75% and 20% of all SC diagnoses, respectively. NMSC, although less lethal, is 18 to 20 times common than CM. The mortality rate is 2 in 1 million. Furthermore, NMSC puts a significant economic burden on health systems, since treatment costs are high, about seven times higher than for CM [4,5] treatment.

By contrast, sun exposure also has beneficial health effects, like the synthesis of vitamin D, which can prevent some types of cancer, and improve mental health and psychological well-being [6,7]. However, it is incorrect to suggest that longer sun exposure will accelerate vitamin D production in regions with high incidences of solar radiation. The UVR levels in tropical regions are sufficiently high for vitamin D to be produced over short, daily exposures, during most parts of the year [8].

Sun blockers like sunscreen creams, sunglasses, long-sleeved clothing, and caps or hats, are all essential for effective protection [9,10,11]. However, daily photoprotection (PhP) also depends on one’s education on the matter. Educational campaigns are essential in combating the harmful effects of UVR and should be supported by behavioral pattern studies on habits and ways receiving sun exposure [12,13,14]. Still, there are not many educational programs on PhP, and a few successful examples can be cited e.g., SunSmart (Australia [15], New Zealand [16]), Sun Safe Schools (UK) [17], SolSano (Spain) [18], and SunWise (United States) [19]. Without proper education on the matter, PhP is normally associated with leisure activities at beaches, pools, tourist locations, sporting activities, or prolonged sun exposure, and most programs are emphasized in the summer. In regions with high UVR levels, everyday exposure can lead to long-term damage, and in general, these educational programs do not address PhP as a necessary daily habit throughout the year.

Recent studies have shown that, regardless of age, profession, and income, PhP habits among individuals are inadequate, even in more developed countries with more robust educational systems [20,21,22,23]. By contrast, the cosmetics industry has reported increased sales of sun creams, which may be related to aesthetic factors related to excessive exposure to the sun, e.g., photoaging, freckles, and wrinkles [24]. Increased PhP use does not always reflect in the proper use of sun creams, sunglasses (effective UV protection), or the use of appropriate clothing or hats that cover parts of the body most prone to the development of SC, like the, lips, eyelids, neck, and extremities [25,26].

There are quite a few studies on PhP use and behavior. However, uncertainties and gaps in knowledge still exist because too few studies have been carried out for equatorial regions with mixed populations. Furthermore, studies that use large sample groups, with more robust statistical tests, are even rarer. These types of studies could allow researchers to assess the influence of other factors, like phototype, age, income, and gender, which are necessary for developing educational policies on PhP, especially in developing countries or countries with high UVR levels throughout the year, e.g., Brazil.

In light of the lack of studies on the topic, this study seeks to: i) detail the increased rates of SC worldwide; ii) demonstrate that UVR levels are higher in tropical climate countries; and, iii) show that PhP measures have been inadequately adopted and that there is a lack of educational policies to mitigate this situation.

The aim of this study was to carry out a detailed assessment of PhP habits and educational programs for children and adolescents in Brazil. The study was carried out with students aged between 4 and 20 years, to relate habits and behaviors among different phototypes, age groups, income levels, and genders. Despite the fact that this study was carried out at the local scale, it should serve as a basis for developing educational prevention campaigns aimed especially at children and adolescents in countries with similar population and economic demographics, and geographical characteristics, in addition to providing subsidies for strategies to map and mitigate the growing numbers of new SC cases.

## 2. Data and Methodology

### 2.1 Sample Characteristics

The study was carried out with children and adolescents aged between 4 and 20 years from four schools in Itajubá (latitude: 22.4S; longitude: 46.6W; altitude: 850m). Three schools were public system schools, and one school was a private school. Itajubá is located in the southeastern region of Brazil and has socioeconomic and educational characteristics similar to those commonly found among the different regions in the country. The sample groups comprised kindergarten, elementary, high school, and college prep students in different stages of study. Itajubá has a typical monsoon climate, with two well-defined seasons, the autumn/winter, from mid-April to mid-September, with cold dry weather, and the while spring/summer, from mid-September to April, with hot and humid weather [27]. Here we adopted a simplified nomenclature referring to these periods as the ‘cold season’ and the ‘hot season’, respectively.

### 2.2. Clothing

In the first stage we conducted visual analysis by taking photographs of the clothing being used, like short or long-sleeved cloths, pants or shorts, open or closed shoes, caps or hats, and accessories and sunglasses. This visual analysis of the clothing was based on an adapted [28] method, and was carried out in two phases, first between March and April (hot season), and second in July (cold season), in 2016. The samples numbered 2162 and 2119 students, respectively, totaling about 4300 students in the two periods. We made observations at the beginning of school (7am and 13pm) and at the end of classes (12pm and 17pm), respectively.

Questionnaires were given to the student that addressed socioeconomic issues [29], a self-assessment of their phototype [30], and their knowledge and habits on PhP [21,31]. Illiterate children were helped by parents or guardians during meetings held at the schools. 512 questionnaires were valid because they were completed in full without erasures. The following socioeconomic requirements were assessed: 1) Gender (2 options): male (M) or female (F); 2) Age group (3 options): a) 4 to 9 years, b) 10 to 14 years, and c) 15 to 20 years; and, 3) Family income, in terms of the number of minimum monthly salaries^1^ (MW) (5 options): a) < 2, b) 2 to 3, c) 3 to 5, d) 5 to 10, and e) >10. Knowledge on PhP and the effects of UVR was assessed using a questionnaire with 14 questions, divided into 3 blocks: a) Knowledge on UVI and use of sunscreen; b) Habit of using accessories for sun protection; and c) Knowledge about the effects of UVR on health. The questionnaires are available in the supplementary material section available at: https://bit.ly/photoprodata.

### 2.4. Statistical analysis

Variance (ANOVA) was analyzed using a three-factor test, followed by the Tukey test (α = 5%) to verify the influence of gender, income, and age group on PhP behavior and education. The answers to each of the questions in the questionnaire were ranked in the ANOVA test. Data sheet with the answers is available in the supplementary material - https://bit.ly/photoprodata. Ranking ranged from 1 for a correct and/or expected answer, to 6 an incorrect/inappropriate answer, corresponding to the total number of items.

The answers were analyzed by ANOVA using a three-factor arrangement (2 × 5 × 3) with 512 samples (questionnaires), considering sex (2 levels), income (5 levels) and age (3 levels), according to what was presented in section 2.3) at 5% probability. In ANOVA, the null hypothesis states that the factors are independent and that there is no interaction among them (p ≥ 0.05), while the alternative hypothesis states that the factors are dependent and that there is interaction among them (p < 0.05). Subsequently, the averages for each factor were submitted to the Tukey test at α = 5%. In the Tukey test, the null hypothesis is that the averages of each level of the non-different factors are (p ≥ 0.05), and the alternative hypothesis is that the averages are different (p<0.05) [32].

## 3. Results

### 3.1. Wearing clothing

Table 1 shows the frequency with which certain types of cloths are worn by students to protect their heads, upper limbs, lower limbs, in addition to shoes and accessories. Itajubá has two well-defined seasons [27], and it is common wear different cloths in the hot and cold seasons. In general, the upper limbs, including the head, eyes, neck, ears and nose, are not adequately protected in both seasons. In fact, caps, and hats are rarely used by students, which is an additional and economically viable option for protecting against the adverse effects of UVR exposure.

**Table 1.**
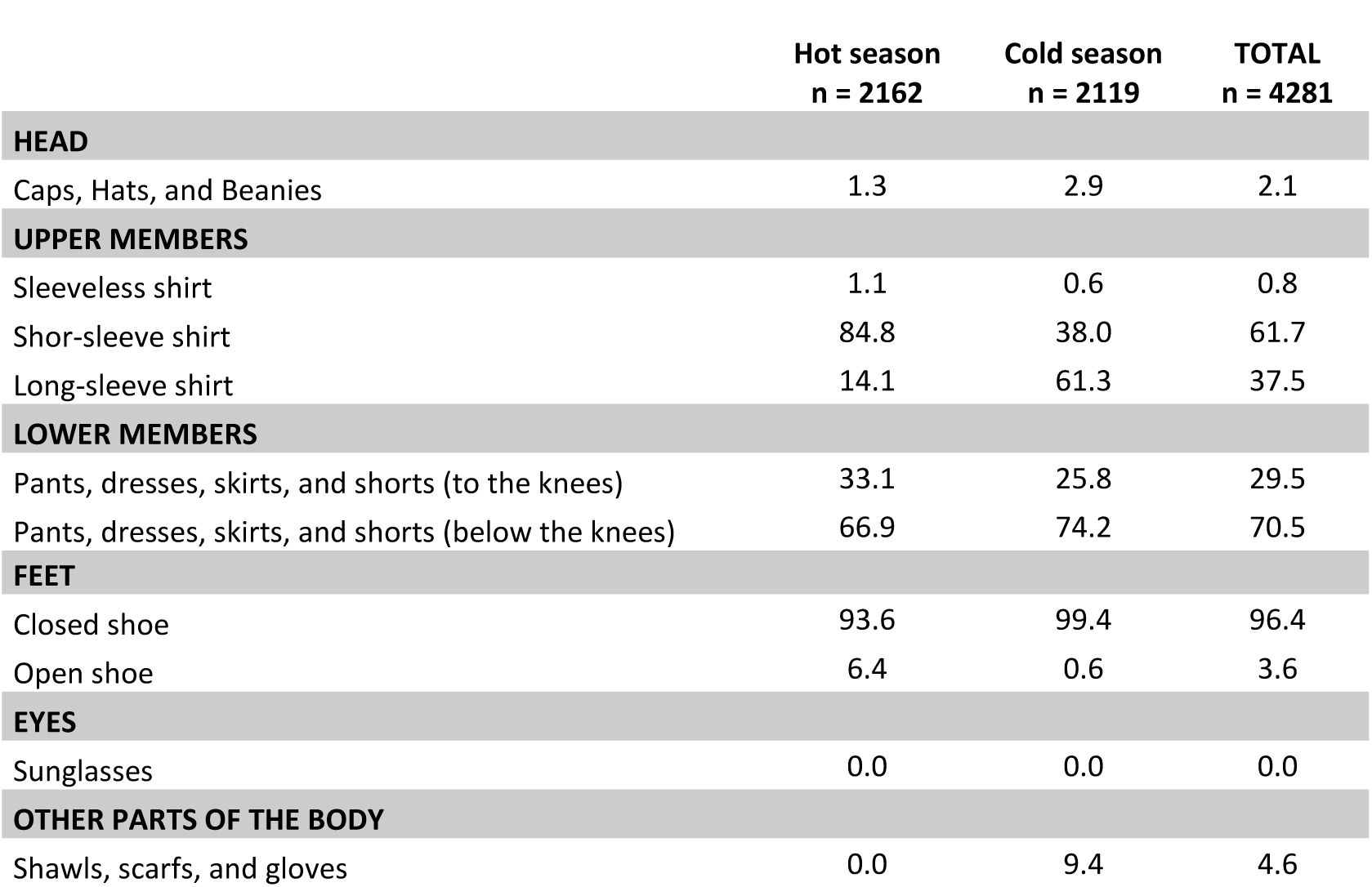
Frequency (%) of wearing PhP clothing and accessories.

|  | Hot season<br>n = 2162 | Cold season<br>n = 2119 | TOTAL<br>n = 4281 |
| --- | --- | --- | --- |
| **HEAD** |  |  |  |
| Caps, Hats, and Beanies | 1.3 | 2.9 | 2.1 |
| **UPPER MEMBERS** |  |  |  |
| Sleeveless shirt | 1.1 | 0.6 | 0.8 |
| Shor-sleeve shirt | 84.8 | 38.0 | 61.7 |
| Long-sleeve shirt | 14.1 | 61.3 | 37.5 |
| **LOWER MEMBERS** |  |  |  |
| Pants, dresses, skirts, and shorts (to the knees) | 33.1 | 25.8 | 29.5 |
| Pants, dresses, skirts, and shorts (below the knees) | 66.9 | 74.2 | 70.5 |
| **FEET** |  |  |  |
| Closed shoe | 93.6 | 99.4 | 96.4 |
| Open shoe | 6.4 | 0.6 | 3.6 |
| **EYES** |  |  |  |
| Sunglasses | 0.0 | 0.0 | 0.0 |
| **OTHER PARTS OF THE BODY** |  |  |  |
| Shawls, scarfs, and gloves | 0.0 | 9.4 | 4.6 |

In general, considering the clothing and accessories that children use, the results show that the upper parts of the body are not adequately protected from the sun in day-to-day life. For example, sunglasses are important for maintaining eye and eyelid health, and yet sunglasses were not used by the students. The students wore short-sleeved clothing, even in the cold season (∼40% of respondents), leaving most of their upper limbs exposed. Accessories like shawls, scarves and gloves, which could provide protection for sensitive regions like the ears, neck, and hands, were only used in winter and among less than 10% of the students.

By contrast, most respondents (between 67% and 74%) preferred to wear pants, dresses, and shorts below the knee line in both seasons, and closed shoes, worn by almost all students (96%), offering adequate protection for the lower extremities of the body.

### 3.2. Socioeconomic aspects and phototypes

Table 2 shows the distribution of phototypes, sex, income, and age. Of the 512 questionnaires applied both sexes were represented equally. Most students were between 10 and 14 years old (46.0%), and showed low-income levels (≤ 2 MW) (57.6%). This distribution is qualitatively close to the sex, age and income distributions for the last census carried out in Brazil [29]. The predominant phototype is type III (> 65% of the sample), followed by phototypes IV (∼17%), and II (∼15%). These results were also compatible with the distribution of skin color in the region, where 76.9% of the individuals state that they are white (phototypes I to III), and 22.7% brown or black (phototypes IV to VI). Since skin color is an individual subjective metric, we can find important variations in this distribution among different regions in Brazil. Comparisons of this nature are complex and beyond the scope of this study.

**Table 2.** Characteristics of the population (sub analysis for gender, age group, family income, and skin phototype) – Relative data in parentheses.

|  |  | Gender |  | Age group (years old)* |  |  | Family income (MW) |  |  |  |  | Skin phototype |  |  |  |  |
| --- | --- | --- | --- | --- | --- | --- | --- | --- | --- | --- | --- | --- | --- | --- | --- | --- |
|  |  | Male | Female | 4-9 | 10-14 | 15-20 | ≤ 2 | 2-3 | 3-5 | 5-10 | > 10 | I | II | III | IV | V |
| **All** | **N = 512** | 260<br>(50.8) | 252 (49.2) | 114<br>(22.3) | 233<br>(45.5) | 165<br>(32.3) | 295<br>(57.6) | 84<br>(16.4) | 84<br>(16.4) | 25<br>(4.9) | 24<br>(4.7) | 1<br>(0.2) | 74<br>(14.5) | 335<br>(65.4) | 88<br>(17.2) | 14<br>(2.7) |
| **Gender** | **Males** | 260<br>(100) | 0 (0) | 57<br>(21.9) | 113<br>(43.5) | 90<br>(34.6) | 139<br>(53.5) | 39<br>(15.0) | 47<br>(18.1) | 17<br>(6.5) | 18<br>(6.9) | 1<br>(0.4) | 27<br>(10.4) | 168<br>(64.6) | 51<br>(19.6) | 13<br>(5.0) |
|  | **Females** | 0 (0) | 252<br>(100) | 57<br>(22.6) | 120<br>(47.6) | 75<br>(29.8) | 156<br>(61.9) | 45<br>(17.9) | 37<br>(14.7) | 8<br>(3.2) | 6<br>(2.4) | 0<br>(0) | 47<br>(18.7) | 167<br>(66.3) | 37<br>(14.7) | 1<br>(0.4) |
| **Age group**<br>**(years old)** | **4-9** | 57<br>(21.9) | 57<br>(22.6) | 114<br>(100) | 0 (0) | 0 (0) | 48<br>(16.3) | 16<br>(19.0) | 18<br>(21.4) | 12<br>(48.0) | 20<br>(83.3) | 1<br>(0.9) | 23<br>(20.2) | 69<br>(60.5) | 17<br>(14.9) | 4<br>(3.5) |
|  | **10-14** | 113<br>(43.5) | 120<br>(47.6) | 0 (0) | 233<br>(100) | 0 (0) | 152<br>(51.5) | 29<br>(34.5) | 40<br>(47.6) | 9<br>(36.0) | 3<br>(12.5) | 0<br>(0) | 32<br>(13.7) | 157<br>(67.4) | 41<br>(17.6) | 3<br>(1.3) |
|  | **15-20** | 90<br>(34.6) | 75<br>(29.8) | 0 (0) | 0 (0) | 165<br>(100) | 95<br>(32.2) | 39<br>(46.4) | 26<br>(31.0) | 4<br>(16.0) | 1<br>(4.2) | 0<br>(0) | 19<br>(11.5) | 109<br>(66.1) | 30<br>(18.2) | 7<br>(4.2) |
| **Family**<br>**income**<br>**(MW)** | **≤ 2** | 139<br>(53.5) | 156 (61.9) | 48<br>(16.3) | 152<br>(51.5) | 95<br>(32.2) | 295<br>(57.6) | 0 (0) | 0 (0) | 0 (0) | 0 (0) | 1<br>(0.3) | 43<br>(14.6) | 190<br>(64.4) | 54<br>(18.3) | 7<br>(2.4) |
|  | **2-3** | 39<br>(15.0) | 45<br>(17.9) | 16<br>(19.0) | 29<br>(34.5) | 39<br>(46.4) | 0 (0) | 84<br>(16.4) | 0 (0) | 0 (0) | 0 (0) | 0<br>(0) | 11<br>(13.1) | 57<br>(67.9) | 13<br>(15.5) | 3<br>(3.6) |
|  | **3-5** | 47<br>(18.1) | 37<br>(14.7) | 18<br>(21.4) | 40<br>(47.6) | 26<br>(31.0) | 0 (0) | 0 (0) | 84<br>(16.4) | 0 (0) | 0 (0) | 0<br>(0) | 14<br>(16.7) | 55<br>(65.5) | 14<br>(16.7) | 1<br>(1.2) |
|  | **5-10** | 17<br>(6.5) | 8<br>(3.2) | 12<br>(48.0) | 9<br>(36.0) | 4<br>(16.0) | 0 (0) | 0 (0) | 0 (0) | 25<br>(4.9) | 0 (0) | 0<br>(0) | 5<br>(20.8) | 14<br>(58.3) | 3<br>(12.5) | 2<br>(8.3) |
|  | **> 10** | 18<br>(6.9) | 6<br>(2.4) | 20<br>(83.3) | 3<br>(12.5) | 1<br>(4.2) | 0 (0) | 0 (0) | 0 (0) | 0 (0) | 24<br>(4.7) | 0<br>(0) | 1<br>(4.2) | 18<br>(75.0) | 4<br>(16.7) | 1<br>(4.2) |
| **Skin**<br>**phototype** | **I** | 1<br>(0.4) | 0<br>(0) | 1<br>(0.9) | 0<br>(0) | 0<br>(0) | 1<br>(0.3) | 0<br>(0) | 0<br>(0) | 0<br>(0) | 0<br>(0) | 1<br>(100) | 0 (0) | 0 (0) | 0 (0) | 0 (0) |
|  | **II** | 27<br>(10.4) | 47<br>(18.7) | 23<br>(20.2) | 32<br>(13.7) | 19<br>(11.5) | 43<br>(14.6) | 11<br>(13.1) | 14<br>(16.7) | 5<br>(20.8) | 1<br>(4.2) | 0 (0) | 74<br>(100) | 0 (0) | 0 (0) | 0 (0) |
|  | **III** | 168<br>(64.6) | 167<br>(66.3) | 69<br>(60.5) | 157<br>(67.4) | 109<br>(66.1) | 190<br>(64.4) | 57<br>(67.9) | 55<br>(65.5) | 14<br>(58.3) | 18<br>(75.0) | 0 (0) | 0 (0) | 335<br>(100) | 0 (0) | 0 (0) |
|  | **IV** | 51<br>(19.6) | 37<br>(14.7) | 17<br>(14.7) | 41<br>(17.6) | 30<br>(18.2) | 54<br>(18.3) | 13<br>(15.5) | 14<br>(16.7) | 3<br>(12.5) | 4<br>(16.7) | 0 (0) | 0 (0) | 0 (0) | 88<br>(100) | 0 (0) |
|  | **V** | 13<br>(5.0) | 1<br>(0.4) | 4<br>(3.5) | 3<br>(1.3) | 7<br>(4.2) | 7<br>(2.4) | 3<br>(3.6) | 1<br>(1.2) | 2<br>(8.3) | 1<br>(4.2) | 0 (0) | 0 (0) | 0 (0) | 0 (0) | 14<br>(100) |

mean age: 12.1 ± 3.9 (all): 6.1 ± 1.7 (4-9yr) / 12.2 ± 1.3 (10-14yr) / 16.1 ± 1.2 (15-20yr)

### 3.3. PhP education on the health effects of UVR

Table 3 shows the results obtained from the questionnaire on PhP education and the health effects of UVR. For simplicity’s sake, this table is a summary of the main results. Complete and detailed results for each factor (gender, age, income and phototype) are available in supplementary material S1 to S4.

**Table 3.** Questions and answers on knowledge and habits for PhP

Questions 1 to 4 – Knowledge about UVI and using sunscreen – multiple answers
|  |  | ANSWERS (%) |  |  |  |  |  |
| --- | --- | --- | --- | --- | --- | --- | --- |
| QUESTIONS |  | a | b | c | d | e | f |
| **Q1** | Did you read in the newspaper that the UV index forecast is 7? Do you think that this radiation level is:<br>a) Low (5)*;<br>b) Medium (4);<br>c) High (1);<br>d) Very High (2);<br>e) Extreme (3);<br>f) I don't know (6) | 2.5 | 16.2 | 27.3 | 9.6 | 1.6 | 42.8 |
| **Q2** | When do you use sunscreen?<br>a) Whenever I leave the house (1);<br>b) Only when I go outdoors to walk, play sports, etc. (2);<br>c) Only when I go to the beach or pool (3);<br>d) Other situations (4);<br>e) I don't use sunscreen (5) | 17.5 | 15.9 | 49.7 | 4.3 | 12.6 |  |
| **Q3** | How do you use sunscreen?<br>a) I apply it only before exposing myself to the sun (2);<br>b) I apply it only when I start to feel the sun heat my skin (3);<br>c) I apply it before exposing myself to the sun and reapply it while I am exposed (1);<br>d) I apply only when I feel I starting to get burned (4);<br>e) I do not apply sunscreen (5) | 43.3 | 5.3 | 32.9 | 2.4 | 16.1 |  |
| **Q4** | When you use sunscreen, what is the sun protection factor (SPF)?<br>a) I don't use sunscreen (4);<br>b) SPF between 6 to 15 (3);<br>c) SPF between 20 to 30 (2);<br>d) SPF between 30 to 60 (1) | 14.3 | 7.6 | 23.9 | 54.2 |  |  |

**B) Questions 5 to 8 – Using sun protection accessories**
|  |  | YES (1)* | NO (3) | SOMETIMES / I DON'T KNOW (2) |
| --- | --- | --- | --- | --- |
| **Q5** | Do you wear any kind of hats or caps when you are outdoors? | 17.0 | 51.8 | 31.2 |
| **Q6** | Do you wear sunglasses? | 21.1 | 48.4 | 30.5 |
| **Q7** | Are your sunglasses UV rated? | 18.4 | 46.7 | 35.0 |
| **Q8** | Do you wear long-sleeved shirts to protect yourself from the sun? | 6.4 | 77.7 | 15.8 |

| C) Questions 9 to 14 – Knowledge on the health effects of UVR |  |  |  |  |
| --- | --- | --- | --- | --- |
|  |  | YES (3)* | NO (1) | I DON'T KNOW(2) |
| **Q9** | Do you think a tan is beautiful and healthy? | 18.4 | 58.6 | 23.0 |
| **Of all the diseases listed below, which are caused by excessive sun exposure?** |  |  |  |  |
|  |  | YES (3)* | NO (1) | I DON'T KNOW(2) |
| **Q10** | Low immunity | 9.4 | 29.5 | 61.1 |
| **Q11** | Skin cancer | 90.8 | 4.5 | 4.7 |
| **Q12** | Eye diseases (cataracts) | 30.3 | 27.7 | 42.0 |
| **Q13** | Premature aging (wrinkles) | 63.1 | 16.0 | 20.9 |
| **Q14** | Skin blemishes (freckles and spots) | 68.8 | 15.0 | 16.2 |
| *Values in parentheses indicate the rank of the response, ranging from 1 for the expected response to the total number of items for incorrect or inadequate responses. Answers are in %. |  |  |  |  |

The first question (Q1) asked an arbitrary question about what UVI = 7 meant, asking students to cite the potential harm of UVR as per the WHO [12]. The results showed a general lack of knowledge surrounding WHO recommendations that seek to increase public awareness about the risks of excessive UVR exposure and to alert them about the need for adopting protective measures. More than 30% of the answers were considered wrong, and 42.8% did not know what UVI stood for. Only 25% of students correctly answered that UVI = 7 represents a high potential health risk. In questions Q2 to Q4, which addressed the use of sunscreen lotions, we observed that half of the students only use sunscreen when they go to the beach or pool, and less than 20% use sunscreen daily. Only 1/3 of the interviewees reapply sunscreen regularly, while 1/4 of the students do not use sunscreen at all or use it incorrectly. 54.2% of students use sunscreen with SPF > 30, as recommended by the medical societies of dermatology [2,3].

Table 3 (part b) shows the questions and answers on using clothing and accessories for sun protection. Hats, sunglasses, and long-sleeved shirts were not used as a form of protection by 52%, 48% and 78% of the students, respectively. Furthermore, most respondents (15 to 30%) admitted to using accessories only sporadically. These students probably don’t view these accessories as being an effective PhP measure, but rather as an aesthetic accessory. Another interesting point, which again highlights the general lack of knowledge on the topic, is that less than 20% of students admitted to knowing if their glasses had UVR protective lenses. It is important to note that the results of this survey coincide with the photographic assessment in section 3.1, and indicate that these accessories are not regularly used by students when they are outdoors. The last sequence of questions (Table 3c) addressed knowledge on the health effects of UVR. More than 90% of students responded that they knew that there was some association between skin cancer and excessive sun exposure. Premature aging and skin spots were cited by 63 and 69% of all respondents, respectively. The greatest gap in knowledge was related to dissociated skin effects, like diseases of the eye and immunosuppression. Only 30% and <10% of all students made this association, respectively. Another interesting aspect, possibly associated with fashion trends, was that approximately 60% of all students did not think that a tan was a desirable or healthy aesthetic quality.

### 3.4. Statistical analyzes of behaviors related to PhP

Table 4 shows the probability of the F test from the ANOVA analyses, and the average comparison between the variation factors (sex, income, and age) using the Tukey test. For brevity’s sake, the values presented in this table are average rankings for the answers referring to each question in the questionnaire (Q1 to Q14). Ranks that tend to 1.00 indicate answers closer to the correct or expected answer for the question. Only three questions (Q8, Q10 and Q12) were not statistically significant (p ≥ 0.05) for any of the three factors considered in this study (gender, income, and age). Long-sleeved shirts (Q8), lowered defenses against infections (Q10), and visual diseases (cataracts) (Q12), were poorly associated with PhP (Tables 1 and 3). There was also no significant relationship between sex, age, and income in the two questions, and the answers indicated less knowledge about diseases caused from excessive sun exposure sun, immunosuppression (Q10), and eye diseases (Q12). There was interaction between gender and age for questions Q1 and Q4, and between gender and income for Q4 (Table 5).

**Table 4.**
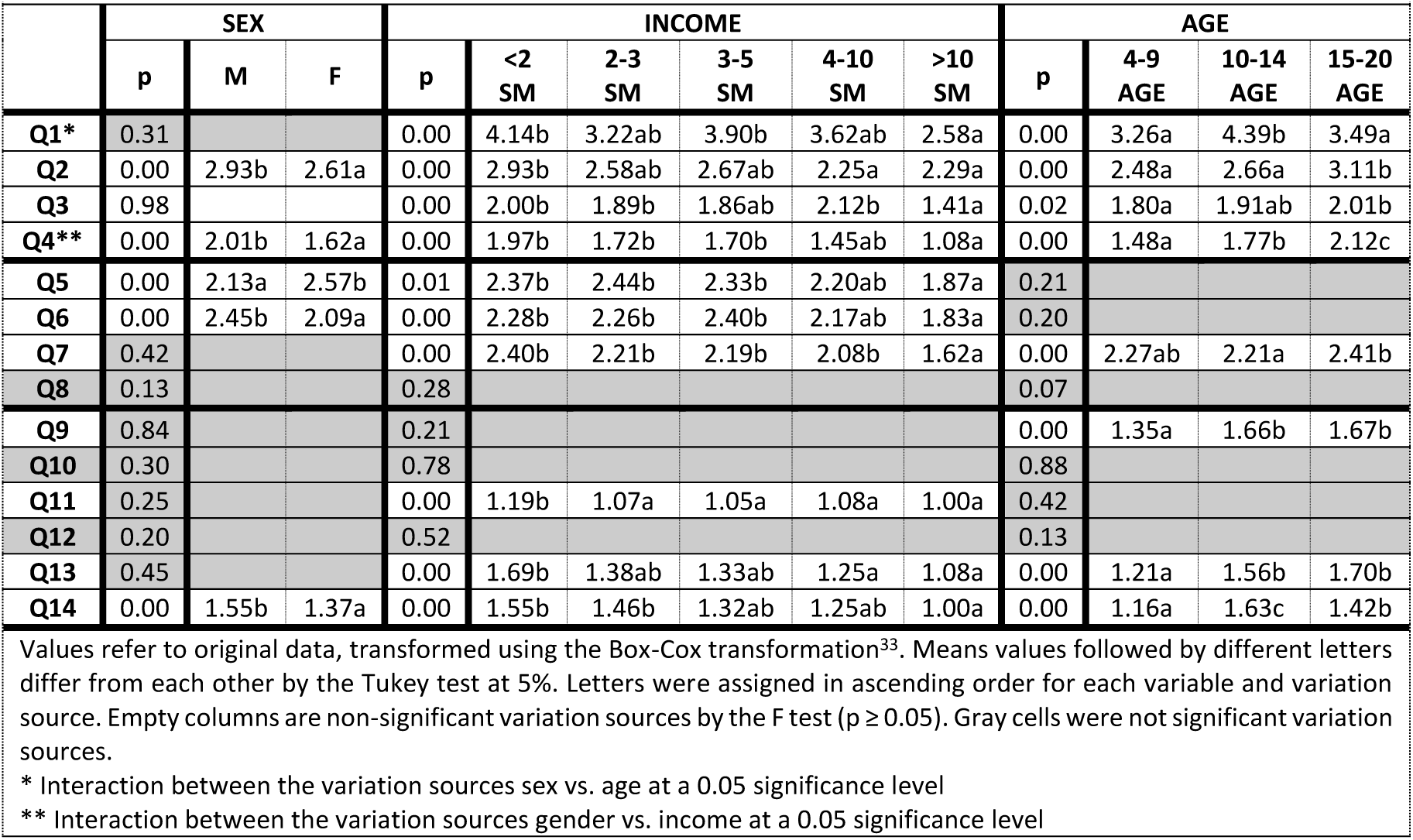
Comparison of the averages for each variable (Q1 to Q14) showing the differences between the levels for each variation factor (gender, income, and age). F-test probability values from the variance analysis for the fourteen questions in Table 3.

|  | SEX |  |  | INCOME |  |  |  |  |  | AGE |  |  |  |
| --- | --- | --- | --- | --- | --- | --- | --- | --- | --- | --- | --- | --- | --- |
|  | p | M | F | p | <2 SM | 2-3 SM | 3-5 SM | 4-10 SM | >10 SM | p | 4-9 AGE | 10-14 AGE | 15-20 AGE |
| Q1* | 0.31 |  |  | 0.00 | 4.14b | 3.22ab | 3.90b | 3.62ab | 2.58a | 0.00 | 3.26a | 4.39b | 3.49a |
| Q2 | 0.00 | 2.93b | 2.61a | 0.00 | 2.93b | 2.58ab | 2.67ab | 2.25a | 2.29a | 0.00 | 2.48a | 2.66a | 3.11b |
| Q3 | 0.98 |  |  | 0.00 | 2.00b | 1.89b | 1.86ab | 2.12b | 1.41a | 0.02 | 1.80a | 1.91ab | 2.01b |
| Q4** | 0.00 | 2.01b | 1.62a | 0.00 | 1.97b | 1.72b | 1.70b | 1.45ab | 1.08a | 0.00 | 1.48a | 1.77b | 2.12c |
| Q5 | 0.00 | 2.13a | 2.57b | 0.01 | 2.37b | 2.44b | 2.33b | 2.20ab | 1.87a | 0.21 |  |  |  |
| Q6 | 0.00 | 2.45b | 2.09a | 0.00 | 2.28b | 2.26b | 2.40b | 2.17ab | 1.83a | 0.20 |  |  |  |
| Q7 | 0.42 |  |  | 0.00 | 2.40b | 2.21b | 2.19b | 2.08b | 1.62a | 0.00 | 2.27ab | 2.21a | 2.41b |
| Q8 | 0.13 |  |  | 0.28 |  |  |  |  |  | 0.07 |  |  |  |
| Q9 | 0.84 |  |  | 0.21 |  |  |  |  |  | 0.00 | 1.35a | 1.66b | 1.67b |
| Q10 | 0.30 |  |  | 0.78 |  |  |  |  |  | 0.88 |  |  |  |
| Q11 | 0.25 |  |  | 0.00 | 1.19b | 1.07a | 1.05a | 1.08a | 1.00a | 0.42 |  |  |  |
| Q12 | 0.20 |  |  | 0.52 |  |  |  |  |  | 0.13 |  |  |  |
| Q13 | 0.45 |  |  | 0.00 | 1.69b | 1.38ab | 1.33ab | 1.25a | 1.08a | 0.00 | 1.21a | 1.56b | 1.70b |
| Q14 | 0.00 | 1.55b | 1.37a | 0.00 | 1.55b | 1.46b | 1.32ab | 1.25ab | 1.00a | 0.00 | 1.16a | 1.63c | 1.42b |

Values refer to original data, transformed using the Box-Cox transformation<sup>33</sup>. Means values followed by different letters differ from each other by the Tukey test at 5%. Letters were assigned in ascending order for each variable and variation source. Empty columns are non-significant variation sources by the F test ( $p \geq 0.05$ ). Gray cells were not significant variation sources.
\* Interaction between the variation sources sex vs. age at a 0.05 significance level
\*\* Interaction between the variation sources gender vs. income at a 0.05 significance level

**Table 5.**
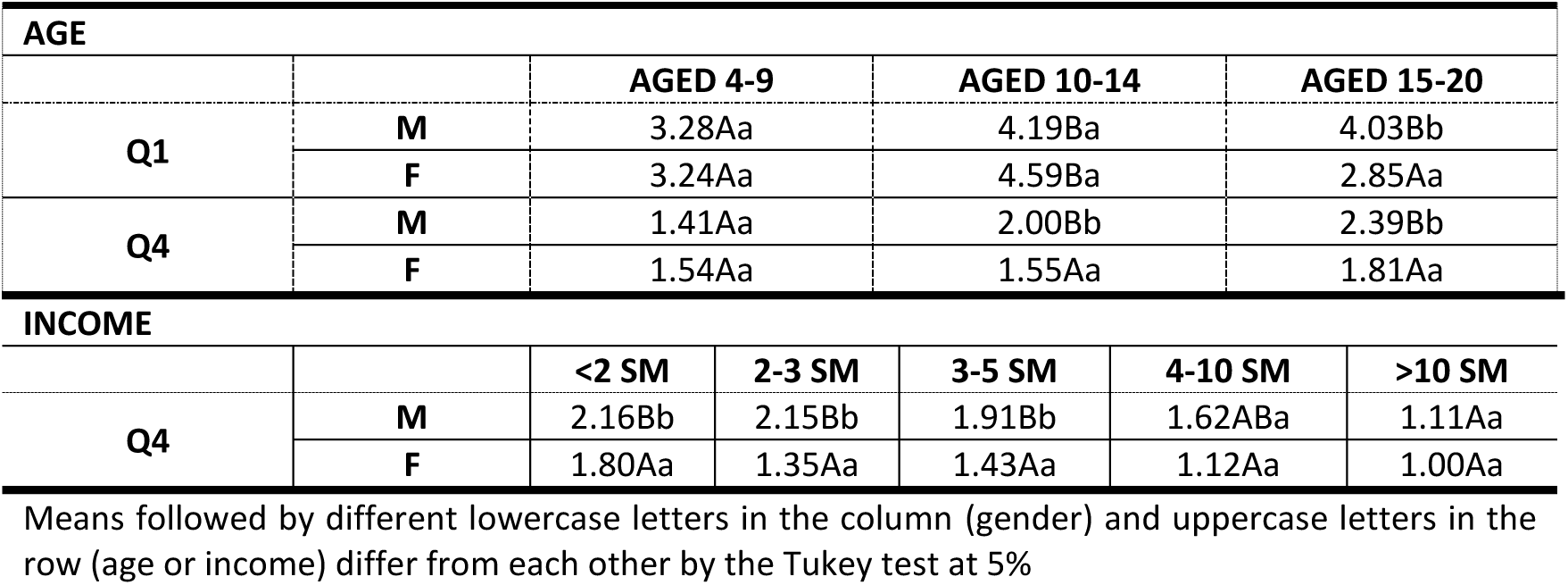
Comparison of the averages for variables Q1 and Q4 that interacted between variation sources (gender vs. age and sex vs. income).

The Income factor showed the greatest number of statistically significant relationships (10) in explaining student knowledge and behavior on PhP (p < 0.05), followed by age (8), and sex (5). The highest income level students (MW > 10) gave answers strongly associated with the expected answers for the three categories of questions presented in Table 3, i.e., the highest income level students sample gave answers closer to the correct and/or expected answers for PhP habits, and for knowledge on UVI and the health effects of UVR.

Although higher income classes had the lowest mean values (ranks) for responses (2.58 for >10 SM, and 3.62 for 4<SM<10), it is important to highlight limitations in knowledge surrounding the meaning of the UVI = 7 scale (Q1). Most responses had high ranks, between 3 and 4, indicating that they partially understood the harmful effect of high UVR exposure levels. Underestimating the harmful effects high UVR exposure levels was corroborated in answers to questions Q2 to Q4 (about the use of sunscreen), Q5, Q6, and Q7 (about using accessories for sun protection). For Q2, the answers had an average rank of 2.65 (±0.28), confirming that sunscreen use is strongly associated with leisure and outdoor activities, regardless of age and gender. Inadequate PhP knowledge was also observed for questions on using accessories for sun protection (Q5-Q8), and was more concerning for lower income classes.

Regarding the age group factor, the answers closest to the correct answer were for younger students (4-9 years). This could reflect parental concerns about PhP in younger children (Q2 to Q4). By contrast, regarding aesthetic tanning (Q9), the results show that students older than 10 were more interested in this aspect. However, most answers did not draw any association between tanning and beauty, thus resulting in a desired prevention aspect.

Finally, the five questions that were significant for gender (Q2, Q4-Q6, and Q14) were answered correctly by female students, except for Q5. Females showed greater concern (and answers closer to the correct answer) regarding questions related to using sunscreen (Q2 and Q4), wearing glasses (Q6), and skin spots, like freckles and moles (Q14). For males, this was true of using accessories like caps and hats (Q5). Using accessories for PhP (Q5 and Q6) was sporadic, and below the desired level for both genders in the questionnaire (rank > 2), and for the on-site observations (Table 1). There was no gender significance for questions on knowledge surrounding the health effects of UVR (Q9 to Q13), except for an association between sun and skin spots.

The relationships between sex, age, and income were also analyzed. There were no statistically significant interactions for the three factors. Interactions between two factors were only observed for questions 1 (gender vs. age), and 4 (sex vs. age and sex vs. income) (Table 5). Similar behavior was observed for both questions in the responses for males and females in younger age groups. By contrast, there was a significant difference between the responses of male and female adolescents (15-20 years). For Q4, there was a significant difference between genders in the responses of groups with MW < 5, as women seemed to understand SPF more than men. For income groups above 5 MW, the responses were homogeneous among both sexes close to the expected/correct values for properly using high SPF sunscreen.

## 4. Discussion and Conclusion

There is little evidence on the effectiveness of photoprotection campaigns for adolescents and adults [34,35]. Primary school may be a critical time for more effective PhP interventions [36,37]. These studies show that there is a lack of evidence on the effectiveness of campaigns, mainly due to gender norms, low sensitivity to the typical risks of adolescent or cultural habits regarding sun exposure. This offers an assessment of the influences of habits, climate, and socioeconomic characteristics to provide greater support and specificity for PhP campaigns. This study was carried out with a statistically significant sample of students, in a place where there is a high incidence of solar radiation and growing numbers of new SC cases diagnosed annually [1]. The studied groups covered age groups from childhood to adolescence mostly characterized by individuals with low purchasing power and phototypes FT ≥ 3.

Hats, sunglasses, and clothing that cover the upper and lower limbs are important physical sun blockers. This study showed these garments are used differently in Brazil than in France [20]. This may be due to the fact that short and long clothes are used in different climates. In southeastern Brazil, winter is predominantly dry with plenty of sunshine. The temperatures are mild, below 22°C, as shown in Figure 1a. In the cold season, slight thermal discomfort was only observed in the early hours of the morning when classes start. The thermal sensation is pleasant during most of the day. For this reason, lighter and shorter clothing is common even during the winter. During the hot season, the thermal sensation is moderately high, and in December, January, and February, high thermal discomfort is common because temperatures are often above 28°C. There is frequent rainfall in this season. These characteristics lead people to use short clothes that exposure the body to more sunlight during most of the year.

**Figure 1:**
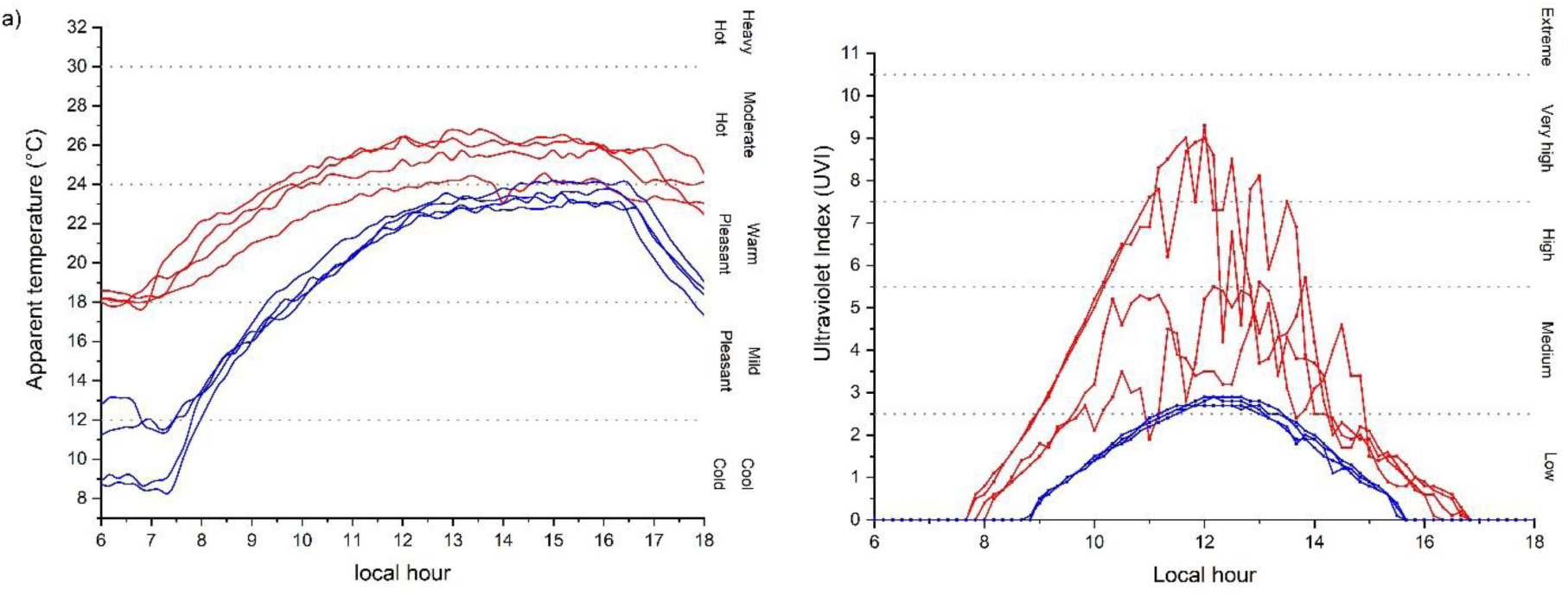
a) Apparent temperature; and, b) Ultraviolet Index during the observational period. Red curves – hot season (March/April 2016); Blue curves – cold season (July 2016)

This aggravates health problems caused by daily exposure to UVR. The UVI data shown in Figure 1b reinforces this point. During the warm period, between 9am and 3pm, about 65% of the UVI measurements are “high” (UVI > 6). About 30% are very high (UVI), and 13.5% are extreme (UVI). According to WHO recommendations, unprotected exposure under these conditions can cause severe damage to the health of the skin and eyes [12]. Typical clear sky days result in relevant UVI levels, even in the winter. UVI levels above the medium health risk scale (3 < UVI < 5) were observed in 85% of the measurements, and high levels (6 < UVI < 7) were observed in approximately 5% of them. According to the WHO, people must protect themselves from the sun under these conditions. The results observed in this study are similar to those observed in other studies carried out in [1,8,38], and corroborate the need for implementing effective educational campaigns into school curricula.

The assessment of PhP education, habits, and knowledge on the nature of UVR and its effects was unsatisfactory, since students showed a profound lack of knowledge about these topics. The UVI is not well known, even though is the main tool for publicizing information on the harmful health effects of the sun. For this reason, information on UVI must be more widely disseminated, and current methods that use media are inefficient. Physical sun blockers use was lower than expected, perhaps due to the local climate that favors short clothes, but also due to the lack of knowledge about the importance of the protection offered by long clothes, hats, and sunglasses. Sunscreen creams are also little used, and are basically only used during leisure activities. Sunscreen is used daily by only 1/5 of all students. When it is used, sunscreen cream is often applied inadequately. In addition to a lack of a culture of awareness surrounding skin protection, one other variable may be the high cost of products. It is worth noting that, despite the lack of sun protection habits, most students knew about the main harmful effects caused by sun overexposure, i.e., skin cancer and aging.

ANOVA allowed us to assess the relationships among the gender, income, and age of respondents. The main result was that higher family income levels were associated with greater student knowledge on the evaluated questions. Younger age groups more adequately used PhP, even though it was still less than ideal. This may be due to the concerns of teachers and parents about the health of the children. Females also more adequately used PhP, possibly because of their concern with aesthetics. It is worth noting that the differences between sunscreen use habits among students of both sexes tend to increase with decreased wage incomes, i.e., teenagers of both sexes with higher incomes had similar concerns about PhP, while sunscreen creams use was higher for women in lower income groups. Recent studies show that tanning as an aesthetic factor in young people may be related to relationship or fashion factors, while for adults, concerns over premature aging discourages aesthetic tanning [39].

Based on the results of this study, we recommended that PhP campaigns take cultural habits, geographic characteristics, and specific socioeconomic aspects into account for the places where these campaigns will be applied. Merely disclosing information about UVI, or simply recommending that people use sunscreen, without educational support, is inefficient in changing the behavior of students, and consequently, mitigating skin cancer. A culture of knowledge surrounding sun protection should be part of education curricula starting in primary school, since there were significant lacks of knowledge on the subject for children and adolescents. Furthermore, campaigns should broaden interest in PhP beyond factors related to aesthetics, more comprehensively encompassing issues related to public health, and incorporating cheaper accessories (like caps, hats, and scarves). This is the only way to broaden interest among low-income groups and males. Finally, we recommend that regional interdisciplinarity PhP policies be developed involving medical, social, economic, and educational experts.

## Data Availability

All data produced are available online at https://bit.ly/photoprodata

## Acknowledgments

We are extremely grateful to all the students who took part in this study. Dr. Corrêa thanks Conselho Nacional de Desenvolvimento Científico e Tecnológico [CNPq, grant 309358/2020-5]. The English grammar review of this article was funded by the Natural Resources Institute of the Federal University of Itajubá. This study was financed in part by the Coordenação de Aperfeiçoamento de Pessoal de Nível Superior – Brasil (CAPES) – Finance Code 001.

## Declarations

### Ethics

This study was approved by the Research Ethics Committee (CEP) of the Faculty of Medicine of Itajubá – FMIt and the National Research Ethics Commission (CONEP).

### Informed Consent

Informed consent was obtained from all participants individually within this study.

### Author contributions

All authors contributed to the study conception and design. Material preparation and data collection were performed by Natana Batista Paiva, Luiz Felipe Silva, Reynaldo José Sant’Anna Silva, Mayara Motta Melo and Clarissa Santos de Carvalho Ribeiro. Data analysis was performed by Marcelo de Paula Corrêa, Fabrina Bolzan Martins and Ana Letícia Campos Yamamoto. The first draft of the manuscript was written by Marcelo de Paula Corrêa and all authors commented on previous versions of the manuscript. All authors read and approved the final manuscript.

## Footnotes

1 One minimum monthly salary in Brazil is a government defined minimum wage for one worker, defined in terms of a monthly salary, NOT an hourly rate. In 2021 the minimum monthly salary in Brazil was R$1,100.00. At the exchange rate USD$1.00 = BRL$5.25 (Aug 29, 2021) this is approximately USD$210.00 a month.

